# What’s UPDOG? A novel tool for trans-ancestral polygenic score prediction

**DOI:** 10.1101/2023.05.15.23289985

**Authors:** David M. Howard, Oliver Pain, Alexandra C. Gillett, Evangelos Vassos, Cathryn M. Lewis

**Affiliations:** Social, Genetic and Developmental Psychiatry Centre, Institute of Psychiatry, Psychology & Neuroscience, King’s College London, UK; Maurice Wohl Clinical Neuroscience Institute, Department of Basic and Clinical Neuroscience, Institute of Psychiatry, Psychology and Neuroscience, King’s College London, London, UK; Department of Medical & Molecular Genetics, Faculty of Life Sciences and Medicine, King’s College London, London, UK

**Author notes:** Corresponding author: David M. Howard, Social, Genetic and Developmental Psychiatry Centre, Institute of Psychiatry, Psychology & Neuroscience, King’s College London, UK.

## Abstract

Polygenic scores provide an indication of an individual’s genetic propensity for a trait within a test population. These scores are calculated using results from genetic analysis conducted in discovery populations. However, when the test and discovery populations have different ancestries, predictions are less accurate. As many genetic analyses are conducted using European populations, this hinders the potential for making predictions in many of the underrepresented populations in research. To address this, UP and Downstream Genetic scoring (UPDOG) was developed to consider the genetic architecture of both the discovery and test cohorts before calculating polygenic scores. UPDOG was tested across four ancestries and six phenotypes and benchmarked against five existing tools for polygenic scoring. In approximately two-thirds of cases UPDOG improved trans-ancestral prediction, although the increases were small. Maximising the efficacy of polygenic scores and extending it to the global population is crucial for delivering personalised medicine and universal healthcare equality.

## Introduction

One of the most promising developments from the field of basic human genetics research has been polygenic scores (PGS)^1^. Large scale genome-wide association studies (GWAS) have enabled the prediction of effect sizes for genetic variants across many different phenotypes. These effect sizes can then be used to calculate an individual’s genetic propensity for a given trait or disease within a population, which is referred to as their PGS^2, 3^. At a population level, the overall prediction of a trait can be estimated using regression models to calculate the total phenotypic variance of a trait that can explained by the PGS in that population^4, 5^.

The theoretical upper bound for the prediction of a trait is its heritability calculated from genomic data^6^. However, the variance explained by PGS are often someway short of that upper bound. There are a number of potential reasons for this shortfall, including non-additive genetic effects^7^, causal variation not well captured by the available variants, differences in measurement of complex phenotypes^8^, differing genetic architectures^9^ and allele frequencies^10^ between populations, and reduced accuracy of effect size prediction from underpowered studies^11^. The differing genetic architecture between ancestries was highlighted by Martin et al. ^12^ as a key reason for lower predictive performance of PGS in non-European ancestry individuals. This lower predictive performance has the potential to further exacerbating global health inequalities due to the current reliance on European samples for conducting GWAS.

One potential solution for improving prediction of PGS across ancestries is to examine the genetic concordance between the GWAS discovery and test populations being examined. If the loci of interest contain a similar genetic architecture due to linkage disequilibrium (LD), then there can be greater confidence in the estimated effect sizes being consistent across ancestries at that position. However, among individuals that have a notably different genetic architecture, the confidence in the consistency of effect size is reduced and could be down weighted to reflect that uncertainty. This assumption led to the development of a software tool known as UP and Downstream Genetic scoring (UPDOG).

The predictive performance of UPDOG was assessed using the GWAS results of six phenotypes from primarily European ancestries and used to calculate and examine prediction in African, East Asian, and South American subsets of the UK Biobank. Genetic variant effect size estimates were obtained for each phenotype using five state-of-the-art PGS tools: Deterministic Bayesian Sparse Linear Mixed Model (DBSLMM)^13^, lassosum^14^, LDPred2^15^, MegaPRS^16^ and PRS-CS^17^. Each tool therefore provided a benchmark for predictive performance that could then be used to examine whether UPDOG improved upon those benchmark predictions.

## Method

Three different models (see Figure 1) and a range of weightings were tested to examine how incorporating genetic data from both the summary statistics, a LD reference panel, and individuals in the test cohort could improve prediction accuracy of PGS. GWAS summary statistics and a LD reference panel were used to examine the effect sizes and LD structure around lead variants in those discovery cohorts. Lead variants are classified as those assigned an effect size by the state-of-the-art PGS tools. Where the LD structure is similar between the discovery cohort and an individual in the test cohort the expectation is that the lead variant’s effect size will have greater accuracy compared to an individual in the test cohort where the LD structure is different. Model A applied adjustments to an individual’s score based on what was carried at the up or downstream position regardless of what was carried at the lead position. Model B applied adjustments where the number of causal or protective alleles at the up or downstream position was different to what was carried at the lead position. Model C applied a haplotype-based approach where the minimum number of causal alleles across the downstream, lead, and upstream position was used. Model B with a scaling factor (*λ*) of 0.025 was identified as the optimum UK Biobank-tuned model (hereafter referred to as UPDOG) and is described below, with Model A and C covered in the supplementary material.

**Figure 1.**
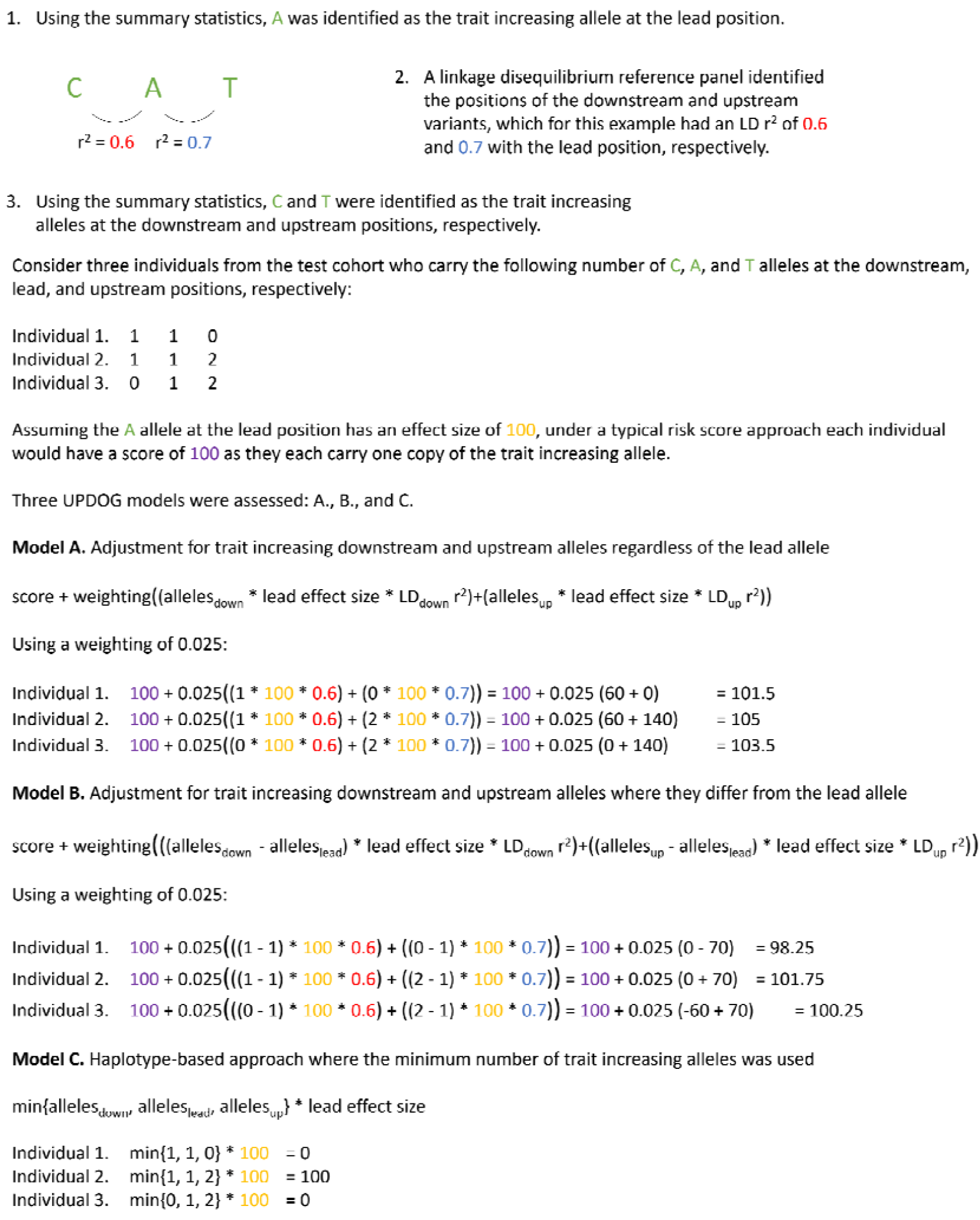
Worked example of the three models that were examined to identify the optimum method for obtaining the greatest number of improvements in trans-ancestral prediction

### UPDOG

UPDOG requires four data inputs 1) the estimated effect sizes of the lead variants for a trait (these can be obtained from *P*-value thresholding and clumping, although here the more advanced methods that incorporate Bayesian or frequentist shrinkage methods are used); 2) the summary statistics providing the genome-wide results from the association analysis of that trait; 3) a LD reference panel matched to those summary statistics, for example, from the 1000 Genome Project^18^ as used here; and 4) the test data for calculating the PGS for individuals in that dataset.

The first step undertaken by UPDOG is to split the genome up into chunks which are then parallelised to reduce the computational burden in terms of both memory and runtime. If estimated effect sizes for over 100,000 genome-wide lead variants are provided, then the genome is split into 1,000 variant chunks, otherwise the genome is split into 30 Mb chunks. LD reference panel data and test data are then created for each chunk and extended by 250 Kb downstream and upstream.

Within each chunk, each lead variant is examined in turn and checked for a match to both the A1 and A2 allele in the LD reference panel data and test data. A downstream variant is then identified using an iterative process moving one variant at a time away from the lead variant. The limits of the LD r^2^ of both the downstream and upstream variants with the lead variant was set between 0.5 and 0.75 within a 250kb window. Therefore, the first variant identified with an LD r^2^ value > 0.5 and < 0.75 and within 250kb of the lead variant is selected as the downstream variant. An upstream variant is identified in the same manner moving in the opposite direction away from the lead variant. An upstream variant is selected if it has an LD r^2^ value > 0.5 and < 0.75, is within 250kb of the lead variant, and has an LD r^2^ value < 0.9 with the selected downstream variant. If the r^2^ value is ≥ 0.9 between the downstream and upstream variant, then the iterative process continues searching for an upstream variant that meets the criteria. The additional stipulation of an LD r^2^ value < 0.9 between the downstream and upstream variants ensures that they both positions provide additional information for the calculation of the PGS.

The calculation of a *score* for each lead genetic variant in the test data follows the standard approach, multiplying the *lead effect size* by the number of alleles carried by an individual at that position^2^. This *score* is then adjusted by examining the difference in the number of *alleles carried* at the *down* and *up* stream positions compared to the number of *alleles carried* at the *lead* variant position with the same direction of effect as the lead variant to produce an UPDDOG score such that:

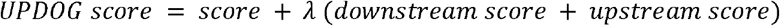

with

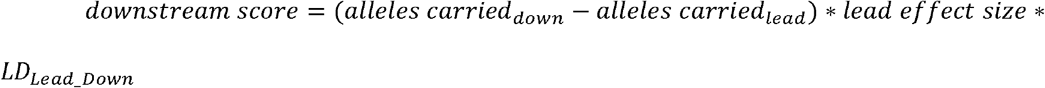

and

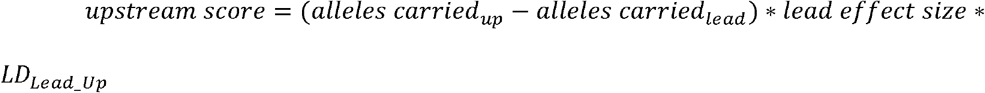

where *LD*_*Lead _Down*_ is the LD r^2^ value between the lead variant and the downstream variant and *LD*_*Lead_Up*_ is the LD r^2^ value between the lead variant and the upstream variant. LD r^2^ value is calculated using the LD reference panel. *λ* is a scaling factor with values of 0.005, 0.0125, 0.025, 0.05, 0.125, 0.25, and 0.5 examined. The *UPDOG scores* are then summed across all lead genetic variants to produce a PGS for each individual with the output from UPDOG being a vector of PGS for the individuals in the test data.

UPDOG is available for download from GitHub: https://github.com/davemhoward/updog

### Evaluation of UPDOG

To evaluate the performance of UPDOG, predictive ability was compared against the benchmark predictions achieved from the five tools used to estimate variant effect sizes across six phenotypes and four different ancestries. The three models (A, B and C) and seven scaling factors (*λ*, as shown in the previous paragraph) were examined to identify the optimum UK Biobank-tuned model.

### Phenotypes and variant effect size estimation

Large genome-wide association studies of six phenotypes were used to examine the predictive performance of UPDOG: body mass index^19^, coronary artery disease^20^, height^21^, major depression^22^, rheumatoid arthritis^23^, and type 2 diabetes^24^. The GWAS for the six phenotypes were selected due to UK Biobank not being included in each analysis (or a version excluding UK Biobank being available in the case of major depression), enabling UK Biobank to be used as the test cohort for PGS prediction. The estimated effect sizes of lead variants for each phenotype, the quality control applied, and the identification of the optimal shrinkage parameters to for obtain effect sizes were the same as those reported in Pain et al. ^25^. Based on the results observed in that paper the estimated effects sizes from DBSLMM^13^, lassosum^14^, LDPred2^15^, MegaPRS^16^, and PRS-CS^17^ were used.

### Testing of prediction in UK Biobank

The UK Biobank is a large health study of over a half a million individuals residing in the United Kingdom^26^. Quality control was applied to the entire UK Biobank study to exclude individuals that had a variant call rate <98%, where the reported sex mismatched the genotypic sex, or where there was relatedness up to the third degree based on a kinship coefficient >0.044 according to the KING toolset^27^. Genetic variants with a call rate⍰<⍰98%, a minor allele frequency <0.01, a deviation from Hardy–Weinberg equilibrium (P⍰<⍰10^−6^), that were non-biallelic, or had an imputation accuracy (Info) score⍰< ⍰0.7 were excluded. This left a total of 7,718,731 genetic variants for the PGS analysis.

The approach described by Privé ^28^ was used to identify 2,161 individuals of African ancestry, 441 individuals of South American ancestry, 1,424 individuals of East Asian ancestry, and a European ancestry subset of 5,913 individuals. The European ancestry subset was restricted to a random sample of 2,000 individuals with ancestry from the United Kingdom and 2,000 individuals with Irish ancestry with the remaining 1,913 individuals with ancestry from Europe (South West), Europe (North East), Ashkenazi, and Finland.

Six phenotypes were assessed in the UK Biobank that were matched to the summary statistics previously described. Body mass index was obtained from Data-Field f.21001.0.0 with individuals that were three standard deviations from the mean removed. Coronary artery disease was based on the definition used by Fürtjes et al. ^29^ incorporating both electronic health records and self-reporting of coronary artery disease to identify cases. Height was obtained from Data-Field f.50.0.0 with individuals that were three standard deviations from the mean removed. For depression, the broad depression phenotype based on help-seeking behaviour from Howard et al. ^30^ was used. Rheumatoid arthritis was based on the respective possible definition used by Glanville et al. ^31^ based on electronic health records and self-report, but without the assessment of medications. Finally, type 2 diabetes was based on the definition used by Fürtjes et al. ^29^, except that an exclusion criteria based on starting insulin within one year diagnosis was not applied (Data-Field f.2986.0.0).

### Assessment of UPDOG

The predictive performance of the respective PGS calculated by UPDOG for the two continuous phenotypes, body mass index and height, was assessed using Pearson’s correlation coefficient. Sex and the first 16 genetic principal components were fitted as fixed effects and the PGS were standardised prior to the calculation of the correlation coefficient. A comparison of the correlation coefficients was made between the original PGS from each state-of-the-art tool and the PGS after applying UPDOG using the same effect sizes from each tool. Comparisons were made for each phenotype and within each ancestry group.

The predictive performance for the four binary phenotypes, coronary artery disease, major depression, rheumatoid arthritis, and type 2 diabetes, was assessed using a covariate-adjusted area under the receiver operating characteristic curve (AUC) and a semiparametric approach^32^ using the AROC R-package^33^. The covariates adjusted were the same as for the continuous phenotypes: sex and the first 16 genetic principal components. A comparison of the AUC was made between the original PGS from each state-of-the-art tool and the PGS after from each state-of-the-art tool after applying UPDOG using the same effect sizes from each tool.

## Results

Summary statistics from six phenotypes (body mass index, coronary artery disease, height, major depression, rheumatoid arthritis, and type 2 diabetes) based on GWAS of European cohorts were analysed for their prediction into ancestral groups (African, South American, East Asian, and European) within the UK Biobank. Predictions were obtained using PGS derived from five state-of-the-art tools (DBSLMM, lassosum, LDPred2, MegaPRS, and PRS-CS) and three potential models (A, B, and C) compared. Model B (UPDOG) performed the best using *λ*= 0.025 with the predictions for each phenotype, ancestral group, and tool compared to the benchmark prediction shown in Figure 2.

**Figure 2.**
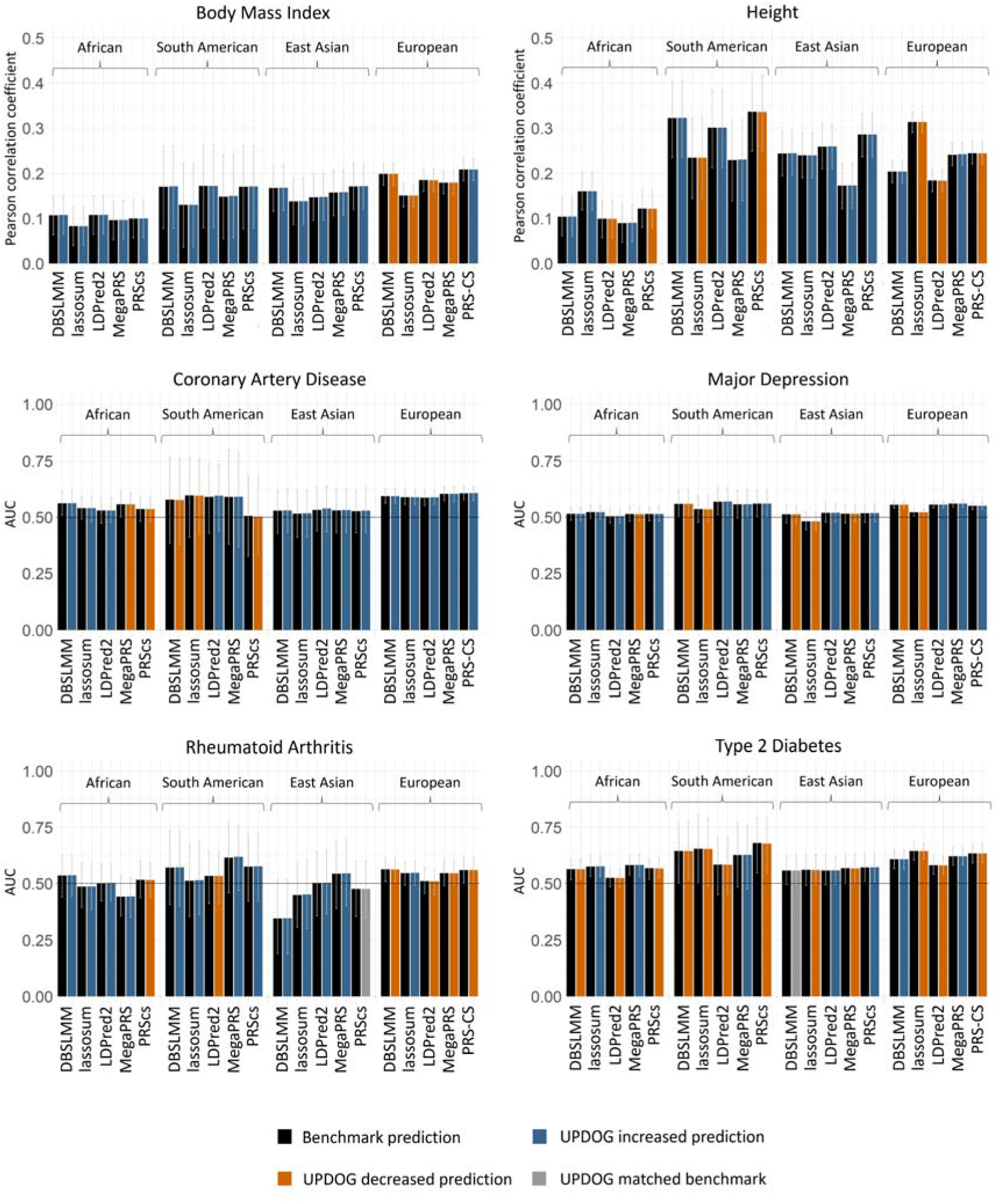
Predictive performance of UPDOG compared to benchmarks set by state-or-the-art tools. Performance was judged across six phenotypes (body mass index, height, coronary artery disease, major depression, rheumatoid arthritis, and type 2 diabetes) using the Pearson correlation coefficient for continuous traits and the area under the receiver operator characteristic curve (AUC) for binary traits. Error bars indicate the 95% confidence interval. Predictions were made in to four ancestries (African, South American, East Asian, and European) using effect size estimates from five state of the art-the-art tools (DBSLMM, lassosum, LDPred2, MegaPRS, and PRS-CS). The effect size estimates from the tools provide a benchmark (shown in black) with which to assess the performance of UPDOG when using those same effect sizes. No change in prediction is shown in grey, increased prediction in blue and decreased prediction in orange.

In total, 62 out of 90 (68.9%) trans-ancestral predictions (based on 6 phenotypes x 5 state-of-the-art tools x 3 ancestries) were improved after applying UPDOG, two (2.2%) were unchanged, and 26 (28.9%) were less predictive. Assuming a binomial distribution, the probability of observing at least 62 improvements out of 90 predictions was 2.2 × 10^−4^.

### Trans-ancestral prediction within each phenotype

For body mass index, all 15 trans-ancestral predictions (3 ancestries and 5 tools) were improved after applying UPDOG using the effects size estimates from each state-of-the-art tool. Eleven out of 15 trans-ancestral predictions for height were improved after applying UPDOG. The average change in the Pearson correlation coefficient for body mass index and height across the 15 predictions was 6.63 × 10^−4^ and 2.07 × 10^−4^, respectively. Coronary artery disease, major depression, rheumatoid arthritis, and type 2 diabetes had improvement in ten, nine, twelve, and five out of 15 trans-ancestral predictions after applying UPDOG, respectively. The average change in the AUC value for coronary artery disease, major depression, rheumatoid arthritis, and type 2 diabetes across the 15 predictions was 8.92 × 10^−4^, 1.35 × 10^−4^, 1.16 × 10^−3^, and -4.04 × 10^−4^, respectively.

### Trans-ancestral prediction within each tool

After using UPDOG to generate polygenic scores from the calculated variant effect sizes from MegaPRS and LDPred2, 14 out of 18 (3 ancestries and 6 phenotypes) predictions improved based on an increased Pearson correlation coefficient or AUC value. The predictions from both DBSLMM and lassosum improved for twelve out of 18 predictions after using UPDOG. The predictions from PRS-CS improved for ten out of 18 predictions after using UPDOG.

### Prediction within each ancestry

Prediction into an African subset from European-based summary statistics was improved for 21 out of 30 (6 phenotypes and 5 tools) predictions after using UPDOG. The prediction into a South American subset and an East Asian subset was improved in 18 and 23 cases out of 30 using UPDOG, respectively. UPDOG was developed to improve trans-ancestral prediction, the analysis within the same ancestry (i.e., prediction from European summary statistics into a European subset in UK Biobank) was also conducted and 14 out of 30 predictions improved after generating polygenic scores with UPDOG.

## Discussion

Polygenic scores are one of the leading scientific advances from the genomic era as they translate genomic findings to individual-level measures of genetic loading^34^. Improving the accuracy and efficacy of PGS to enable their use within clinical settings to deliver personalised treatments is a vital area of research. Three key opportunities exist for improving predictions from PGS. Firstly, to improve the accuracy of the effect size estimates from genome-wide association studies. This can primarily be achieved through increased sample size or increased accuracy in phenotype ascertainment. Second, improving the process for incorporating the genome-wide effect sizes from multiple correlated genetic variants. Primarily this has been through the optimisation of shrinkage methods and lead to the development of state-of-the-art tools such as LDPred2^15^, MegaPRS^16^, and PRS-CS^17^. Finally, developing novel methods to use the calculated effect sizes to determine the PGS assigned to each individual in a test cohort. Where the genetic architecture is similar between the samples used in the genome-wide association study and the test cohort the standard approach of summing the risk alleles weighted by their assigned effect sizes^2^ is likely to be most effective. However, where there are differing genetic architectures, potentially due to ancestry, between the samples used in the genome-wide association study and the test cohort then alternate approaches are required.

UPDOG represents a novel approach for calculating PGS when the effect size estimated for genetic variants are obtained using an ancestry that is different to the one in which the predictions are being made. By examining the genetic architecture surrounding the lead genetic variants based on the summary statistics and comparing those alleles with what each individual carries in the test cohort improvements in prediction should be possible. The performance of UPDOG was examined across four ancestries, six phenotypes, and benchmarked against five state-of-the-art tools for the calculation of variant effect sizes. The number of increases in prediction were above that expected by random chance (*P* = 2.2 × 10^−4^); however, the increases in either Pearson correlation coefficient or AUC value were modest.

The number of improved predictions from UPDOG was relatively consistent using the calculated effect sizes from the five state-of-the-art tools and across the three non-European ancestries. Among the six phenotypes tested, body mass index had the greatest number (15 out of 15) of improved trans-ancestral predictions when applying UPDOG. Rheumatoid arthritis, height, coronary artery disease, and major depression had between 9 and 12 increase trans-ancestral predictions. Type two diabetes was only improved for 5 out of 15 trans-ancestral predictions. It is unclear why there were differences between phenotypes for the number of improved predictions. The use of only UK Biobank participants should help reduce any bias in phenotypic ascertainment between the ancestries analysed compared to ancestries drawn from different cohorts. Power due to genome-wide association study sample size is unlikely to be the cause of these differences, with depression having the largest sample size and rheumatoid arthritis having the smallest. SNP-based heritability (h^2^) may have contributed to the performance of UPDOG with body mass index (h^2^ = 0.49) and height (h^2^ = 0.25) being more heritable than the other phenotypes (0.07 < h^2^ < 0.16) based on estimates from the UK Biobank SNP-Heritability Browser^35^. Body mass index and height were also continuous measures whereas the other phenotypes used binary measures of case control status.

An alternative tool for calculating PGS using data from more than one ancestry is PRS-CSx^36^. PRS-CSx enables the inclusion of results from multiple genome-wide association studies conducted using different ancestries and applies a shared continuous shrinkage prior and models population-specific allele frequencies and LD patterns to produce posterior variant effect size estimates. A multi-ethnic polygenic risk scores approach has also been suggested by Márquez-Luna et al. ^37^ and uses a sample size weighted average of estimated effect sizes to generate variant effect size estimates. Both these approaches seek to maximise the sample size and optimise the output of the calculated effect sizes. Currently UPDOG is the only tool that also considers the individuals within the test cohort. However, UPDOG is not designed to consider either genome-wide association studies that have meta-analysed multiple ancestries or the inclusion of separate ancestry-specific association studies. The results presented here are based on prediction into individuals in the UK Biobank and may not be informative for individuals outside of this setting. The results presented here are potentially biased by the selection of the optimum model and weighting using the same sample for which the results are reported.

Strategies for increasing the predictive performance of PGS for disorders and diseases are crucial in ensuring that the right person gets the right support at the right time. There has very rightly been an extensive effort to widen genotyping to include more individuals from under-represented populations. Maximising the potential of this data is critical for overcoming global health inequalities. Therefore, UPDOG was developed to improve prediction of polygenic scores across different ancestries. UPDOG was able to provide an increase in trans-ancestral prediction in over two-thirds of instances examined using effect sizes estimated from state-of-the-art tools. The improvements in Pearson correlation coefficients or AUC values delivered by UPDOG were small. However, the development of tools that also consider the population being predicted into is an area of research that warrants further investigation.

## Data Availability

The software used in this study is available at: https://github.com/davemhoward/updog
All data produced in the present study are available upon reasonable request to the authors.

## Acknowledgements

This research was conducted using the UK Biobank resource, application number 16577. We are grateful to the UK Biobank and all its voluntary participants. The UK Biobank study was conducted under generic approval from the NHS National Research Ethics Service (approval letter dated June 17, 2011, Ref 11/NW/0382). All participants gave full informed written consent.

D.M.H is supported by a Sir Henry Wellcome Postdoctoral Fellowship (Reference 213674/Z/18/Z). C.M.L acknowledges MRC grant MR/N015746/1. This investigation represents independent research part-funded by the National Institute for Health Research (NIHR) Maudsley Biomedical Research Centre at South London and Maudsley NHS Foundation Trust and King’s College London. The views expressed are those of the authors and not necessarily those of the NHS, the NIHR or the Department of Health and Social Care.

This research was funded in whole, or in part, by the Wellcome Trust [Reference 213674/Z/18/Z]. For the purpose of open access, the author has applied a CC BY public copyright licence to any Author Accepted Manuscript version arising from this submission.

## Authorship Contributions

D. M. H., O. P, A. C. G., E. V., and C. M. L. contributed to the concept and design of the work. D. M. H. developed the new software (UPDOG) used in the work and conducted the analysis. D. M. H., O. P, A. C. G., E. V., and C. M. L. contributed to the interpretation of the results. D. M. H. drafted the manuscript which was subsequently revised by O. P, A. C. G., E. V., and C. M. L.

## Data availability

According to Wellcome Trust’s Policy on data, software and materials management and sharing, all data supporting this study will be openly available at http://doi.org/doi:10.XXXXX/XXXXXXXXX

