## Supplementary Information for "What’s UPDOG? A novel tool for trans-ancestral polygenic score prediction"

Supplementary Material

Three different models were examined. Model B provided the greatest number of increased predictions summed across three ancestries (African, East Asian, and South American), six phenotypes (body mass index, coronary artery disease, height, major depression, rheumatoid arthritis, and type 2 diabetes) and five state-of-the-art tools (DBSLMM, lassosum, LDPred2, MegaPRS, and PRS-CS) and is reported in the main manuscript. The methods for Models A and C and the overall performance from the three models are reported below.

Model A

Model A requires the same four data inputs reported in the main manuscript: the estimated effect sizes of the lead variants for a trait, the summary statistics, a linkage disequilibrium (LD) reference panel matched to those summary statistics and the test data.

The upstream and downstream variants are identified using the same method reported in the main manuscript (LD r^2^ value > 0.5 and < 0.75 and within 250kb of the lead variant, and upstream and downstream variants with an LD r^2^ value < 0.9).

The calculation of a $score$ for each lead genetic variant in the test data follows the standard approach, multiplying the $lead effect size$ by the number of alleles carried by an individual at that position (Choi et al., 2020). This $score$ is then adjusted by examining the number of $alleles carried$ at the $down$ and $up$ stream positions that have the same direction of effect as the lead variant to produce an UPDDOG score such that:

$$UPDOG score = score + \lambda(downstream score + upstream score)$$

with

$downstream score={alleles carried}_{down}*lead effect size*{LD}_{Lead\_Down}$

and

$upstream score={alleles carried}_{up}*lead effect size*{LD}_{Lead\_Up}$

Where $\lambda$ is a scaling factor, ${LD}_{Lead\_Down}$ is the linkage disequilibrium between the lead variant and the downstream variant, and ${LD}_{Lead\_Up}$ is the linkage disequilibrium between the lead variant and the upstream variant. The linkage disequilibrium observed in the LD reference panel matched to those summary statistics is used for ${LD}_{Lead\_Down}$ and ${LD}_{Lead\_Up}$. The $UPDOG scores$ are then summed across all lead genetic variants to produce a PGS for each individual with the output from UPDOG being a vector of PGS for the individuals in the test data.

Model C

Model C requires the same four data inputs reported in the main manuscript: the estimated effect sizes of the lead variants for a trait, the summary statistics, a linkage disequilibrium (LD) reference panel matched to those summary statistics and the test data.

The upstream and downstream variants are identified using the same method reported in the main manuscript (LD r^2^ value > 0.5 and < 0.75 and within 250kb of the lead variant, and upstream and downstream variants with an LD r^2^ value < 0.9).

Model C concurrently examines the alleles at the downstream, lead, and upstream positions to ensure the causal alleles are carried at each position. Across the three positions the minimum number of causal $alleles carried$ are multiplied by the $lead effect size$ to calculate an $UPDOG score$ for each lead genetic variant in the test data:

$$UPDOG score = {min\{alleles carried}_{down},{alleles carried}_{lead},{alleles carried}_{up}\}*lead effect size$$

The $UPDOG scores$ are then summed across all lead genetic variants to produce a PGS for each individual with the output from UPDOG being a vector of PGS for the individuals in the test data.

Model results

The performance of the three models were assessed based on the number of improvements in predictions observed across 90 tests using European summary statistics to predict in to African, East Asian, and South American ancestry subgroups in the UK Biobank (Supplementary Figure 1). Model B with a scaling factor (λ) of 0.025 produced 62 improvements in prediction out of a possible 90 tests and was identified as the optimum model with further analysis provided in the main manuscript.

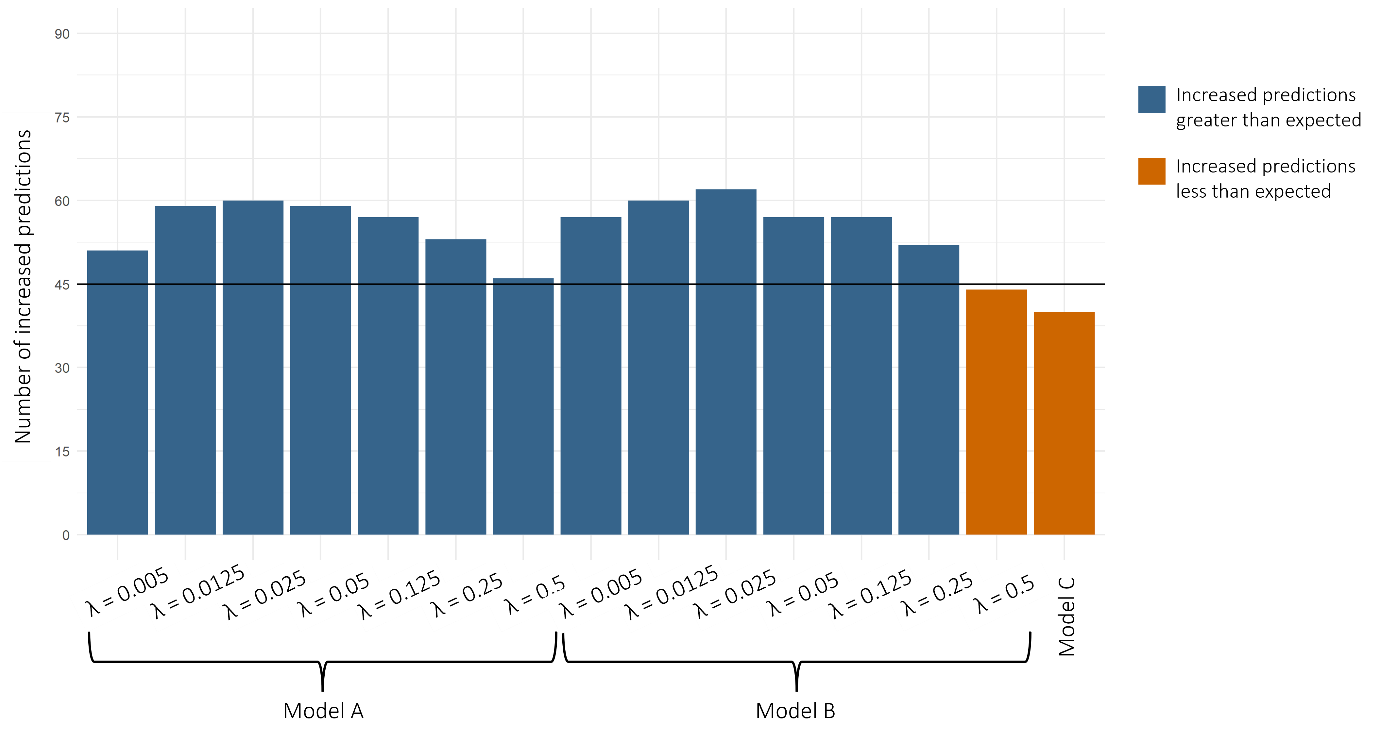

Supplementary Figure 1. The number of predictions that increased after applying the three models. Models A and B use a scaling factor (λ). There were 90 tests conducted and the solid horizontal line represents the expected number of increased predicted by random chance (45 tests). Where the number of increased predictions exceeds the expectation, the bar is coloured blue, else where the number of increased predictions is below that expected the bar is coloured orange.
